# Sympathetic nerve activity recovery from the skin recording using the modern optimal shrinkage technique

**DOI:** 10.1101/2025.01.23.25321036

**Authors:** Pei-Chun Su, Chao-Yi Chen, Chia-Hao Kuo, Wei-Chung Tsai, Hau-Tieng Wu

## Abstract

**Objective:** The widely used bandpass filter (BPF)-based algorithm for recovering sympathetic nerve activity (SNA) from the skin sympathetic nerve activity (SKNA-I) signal, recorded via electrocardiogram electrodes or subcutaneous sympathetic nerve activity (SCNA-I) in a lead I setup, has limitations. It excludes spectral information outside the BPF range and may retain artifacts, such as cardiac activity or pacemaker interference, in the recovered SNA (rSNA) signal. This study aims to develop an algorithm that recovers the full spectral SNA information as comprehensively as possible for evaluating the autonomic nervous system (ANS).

**Methods:** We propose a novel algorithm, S3 (*SNA from Shrink and Subtraction*), which integrates the optimal shrinkage algorithm (eOptShrink) with the template subtraction (TS) method. The performance of S3 was evaluated against other algorithms using semi-real simulated SKNA-I data, a human SKNA-I database including subjects with pacemakers or atrial fibrillation, and a mouse SCNA-I database.

**Results:** The S3 algorithm demonstrated numerical efficiency and outperformed existing approaches, including traditional TS, BPF and other methods, in both time and frequency domains. Notably, in addition to the traditional 500-1000Hz spectral band, S3 effectively recovers spectral information across the 50-300Hz and 300-500Hz frequency bands, making it suitable for homecare ANS evaluation. All quantitative results are supported by the rSNA tracing for visual inspections.

**Conclusion:** S3 accurately recovers the full-spectrum SNA.

**Significance:** By enabling the exploration of the entire SNA spectrum, S3 offers a promising tool for ANS evaluation and applications in homecare environments.

## 1. Introduction

Autonomic nervous system (ANS) dysregulation, particularly sympathetic hyperactivation, is a significant and relevant clinical problem. ANS modulation plays a very important role in arrhythmias [1]. Dysregulation of the sympathetic and parasympathetic nervous systems can lead to arrhythmias, particularly atrial fibrillation (AF) [2] and ventricular arrhythmia (VA). Electrical and mechanical remodeling of the ANS, particularly sympathetic dysregulation following myocardial infarction (MI), induces catecholamine release, which enhances automaticity, triggered activity and reentrant mechanisms of VA [3]. Clinical VA is frequently preceded by sympathetic activation [4]. In addition, sympathetic overdrive after MI can provoke further myocardial ischemia, leading to a vicious circle and sudden cardiac death. Therefore, early detection of sympathetic hyperactivation is crucial for managing high-risk patients.

The stellate ganglion (SG), the most important extracardiac intrathoracic sympathetic ganglion, is associated with cardiac arrhythmias [5, 6]. SG blockade is a key treatments for VA storms [7]. The direct communication between the SG and the cervical and thoracic skin provides a physiological basis for using skin sympathetic nerve activity (SKNA) to estimate stellate ganglion nerve activity (SGNA) in canine models [8, 9] and in humans [10-13], offering an alternative to microneurography for sympathetic nerve activity (SNA) recording in humans. SKNA can be easily recorded using conventional electrocardiogram (ECG) electrodes, making it an ideal clinical approach for evaluating SNA. It has been applied to study atrial fibrillation [13-15], ventricular arrhythmia [12], obstructive sleep apnea [16], prognosis prediction of critically ill patients [17], syncope [18], cognitive stress evaluation [19] and acute coronary syndrome [20]. In addition, subcutaneous sympathetic nerve activity (SCNA) has been used to estimate SGNA in an ambulatory canine model of MI and may provide a better correlation with SGNA than heart rate variability (HRV) [21].

Traditionally, the human SNA signal is estimated in the following way. First, electrical potential is recorded using a pair of bipolar electrodes placed in the right and left subclavian areas following the lead I setup, referred to as SKNA-I, at a high sampling rate, such as 10,000 Hz. The SNA is estimated by applying a BPF between 500 and 1000 Hz [22], while the ECG signal can be recovered by applying a band-pass filter (BPF) between 0.05 Hz and 150 Hz. Hereafter, Similarly, SCNA in animal studies is recorded in a comparable manner, with the electrode placed beneath the skin to directly capture activity from the subcutaneous nerves in a lead I configuration. The signal is referred to as SCNA-I, and the SNA can be estimated by applying the same BPF. Hereafter, we refer to the estimated SNA as the *recovered* SNA (rSNA). If a specific algorithm is used, such as ALGO, we denote it as rSNA-ALGO.

Although the BPF approach is widely used, it faces certain limitations. First, while SNA has been reported to have a spectral range from 0 to 2000 Hz [22], the BPF limits the rSNA to a narrow band, restricting researchers from exploring potential information in other spectral bands. Second, BPF can suffer from spectral leakage. Cardiac activity in the ECG signal is quasi-periodic and characterized by sharp R peaks, resulting in a spectral range primarily composed of harmonics of the P-QRS-T waveform. This range can widen with heart rate variability, allowing cardiac activity to remain in rSNA-BPF. Figure 1 illustrates this with a SCNA-I signal recorded from a mouse with premature ventricular contractions (PVC) in (a) and its filtered version (500-1000 Hz BPF) in (b). This challenge is especially pronounced when studying pediatric SNA, where the spectral range of cardiac activity may span from 0.05 to 250 Hz [22] due to the higher heart rate, or when using ambulatory ECG monitors with a sampling rate at 1000 Hz, where a BPF range of 300 to 500 Hz is recommended [23]. Third, for patients with an active pacemaker, pacing artifacts, which are essentially sharp delta peaks, introduce strong interference in the BPF output. Figure 1 shows this in (d) with an SKNA-I signal with pacing artifacts and in (e) with a filtered (500-1000 Hz) version, where residual artifacts are still evident. In summary, a new algorithm is needed to effectively remove cardiac interference and recover the SNA. Addressing these issues is the focus of this study.

**Figure 1.**
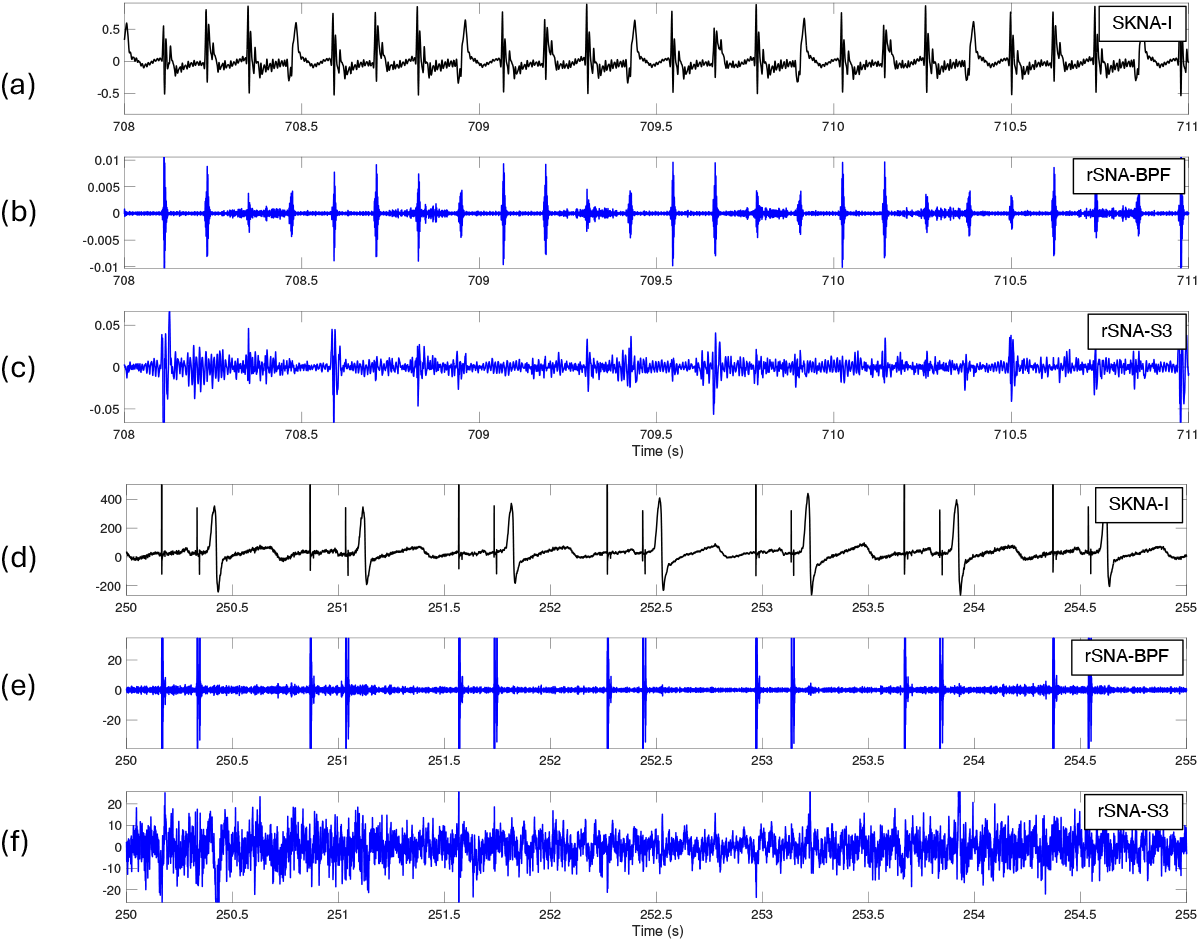
An illustration of the SCNA-I and SKNA-I signals (a and c), the rSNA-BPF with the spectral range 500-1000Hz (b and e), and the rSNA-S3 (c and f).

By observing the periodic pattern in the SKNA-I signal, a straightforward approach to eliminating cardiac activity is the template subtraction (TS) algorithm. This method assumes that the SKNA-I signal consists of a clean ECG signal superimposed with the SNA signal, where each cardiac activity event within the clean ECG follows a highly similar pattern over time. Based on this assumption, by subtracting the average of all or nearby cardiac cycles detected in the SKNA-I signal from the SKNA-I signal, the SNA signal is obtained. While this method has been widely used for applications such as fetal ECG decomposition from transabdominal ECG [24] and stimulation artifact removal from local field potential recordings [25], certain limitations of the TS approach have been noted. Specifically, the assumption of identical or nearly identical repeating cardiac cycles is often invalid due to physiological dynamics. In this study, factors such as respiration and arrhythmias affect the periodicity of cardiac cycles, causing them to vary from beat to beat. To address these challenges, we refine the TS approach by replacing the assumption of identical cycle patterns by the wave-shape manifold structure [26] of ECG and accommodating the nonstationary characteristics of SNA. The process is summarized in 4 steps: detect cardiac cycles, apply the optimal shrinkage (OS) algorithm called eOptShrink [27] to separate SNA from cardiac cycles, stitch the cleaned-up cardiac cycles to recover the ECG, and ultimately recover the SNA. Given the nature of the proposed algorithm, we coin it S3, which stands for *SNA from Shrink and Subtraction*, and call the result rSNA-S3. See Figure 1(c)(f) for the results of the proposed S3 algorithm.

The paper is organized in the following. In Section 2, motivated by physiological insights, we propose a phenomenological model to describe the SKNA-I or SCNA-I signal. In Section 3, we introduce the proposed S3 algorithm in detail. The database and statistical methods are presented in Section 4. Section 5 showcases the numerical results. The paper concludes with a discussion in Section 6 and a final conclusion in Section 7.

## 2. Mathematical model

Consider the SKNA-I recording as the SNA contaminated by cardiac signal, which we call electrocardiogram (ECG) to simplify the discussion. Note that the ECG component in the SKNA-I is quasi-periodic, with not only the amplitude and frequency changing from time to time caused by, for example, respiration and heart rate variability, but also the oscillatory pattern changing from time to time, caused by, for example, QT variability caused ventricular repolarization dynamics or arrhythmia. It is possible that the subject receives pacemaker treatment, and the cardiac cycle contains pacing artifacts representing as delta-like peaks. These facts lead to the following *phenomenological* model. See Figure 2 for an illustration of the proposed model for SKNA-I or SCNA-I.

**Figure 2.**
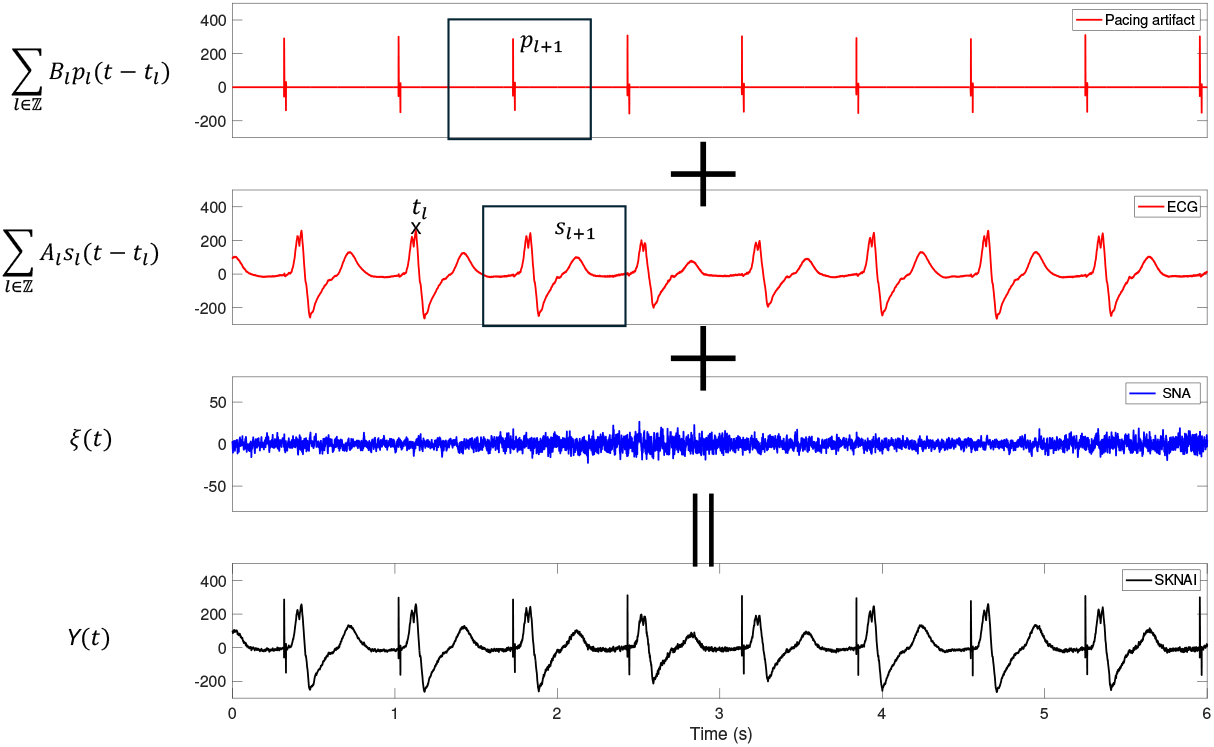
An illustration of the proposed phenomenological model.

Suppose *t*_*l*_ is the timing of the *l*-th *R peak*, so that … < *t*_*l*_ < *t*_*l*+1_ < *t*_*l*+2_ < ⋯. Model the SKNA-I or SCNA-I by a random process

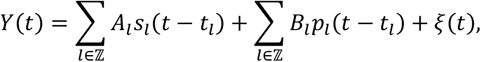

where ∑_*l*∈𝕫_ *A*_*l*_*s*_*l*_(*t* − *t*_*l*_) is deterministic representing the ECG, ∑_*l*∈𝕫_ *B*_*l*_*a*_*l*_(*t* − *t*_*l*_) is deterministic representing the potential pacing artifacts, and ξ(*t*) is stochastic representing the SNA we have interest, which might include electromyogram and inevitable noise. When the pacemaker is installed, we model ECG and pacing artifacts separately to respect their different electrophysiological roles. Given a realization of *Y*(*t*), the signal processing goal is decomposing it into ∑_*l*∈𝕫_ *A*_*l*_*s*_*l*_(*t* − *t*_*l*_), ∑_*l*∈𝕫_ *B*_*l*_*p*_*l*_(*t* − *t*_*l*_) and the realization of ξ(*t*). We now detail this model.

First, *A*_*l*_ > 0 and *B*_*l*_ ≥ 0 describe the magnitude of the *l*th cardiac cycle and the associated pacing artifacts that is impacted by respiration. *s*_*l*_ is a compactly supported smooth function that describes the electrophysiological dynamics of the *l*-th cardiac cycle, including the P wave if the subject is atrial fibrillation (Af) free, QRS complex and T wave. We call *s*_*l*_ the *wave-shape function* (WSF) [28] associated with the *l*th cardiac cycle. Note that the R peak to R peak interval (RRI) time series, *t*_*l*_ − *t*_*l*-1_, is not constant due to heart rate variability, and it exhibits characteristics similar to white noise with a mean of zero during episodes of Af.

When a subject is free of pacemaker treatment, *B*_*l*_ = 0, and we do not have pacing artifacts. In a subject with pacemaker, *B*_*l*_ > 0, and the cardiac cycle includes artifacts caused by the pacing, particularly those delta-like peaks coming from the pacing. Denote the artifact associated with the *i*th cardiac cycle *p*_*i*_, which contains delta-like peaks. The number of peaks in *p*_*i*_ depends on the pacing mode. In the DDD (Dual Chamber Pacing, Dual Chamber Sensing, Dual Response) mode, both inhibit and trigger pacing based on sensed events exist, which leads to 4 delta-like peaks, two before P waves and two before QRS complex. In the VVI (Ventricular Pacing, Ventricular Sensing, Inhibited Response) mode, we have two delta-like peaks before QRS complex. We assume both *s*_*l*_ and *p*_*l*_ have unit *L*^2^ norm an *s*_*l*_(0) is the associated R peak. We call *p*_*l*_ the WSF of its associated pacing artifact if the subject is under the pacemaker treatment.

Clearly, both *s*_*l*_ and *p*_*l*_ have nonrandom patterns and follow electrophysiology rules. Mathematically, each WSF can be modeled to be parametrized by a low dimensional manifold embedded in the sphere of *L*^2^(ℝ), called the wave-shape manifold [26]. We refer readers with interest to [26] for details. This is the key assumption for our algorithm design.

The random process ξ(*t*) models the combined contribution of SNA, myopotential, and noise. Myopotential signals can have frequency components as high as 400 Hz but are generally concentrated below 100 Hz. SNA reflects the electrical activity of sympathetic nerves that innervate sweat glands and blood vessels in the skin, with frequency components ranging from 0 to >2000Hz and amplitudes typically between 0.5 to 100 μV, though they can be higher [22]. SNA serves as an indirect measure of sympathetic nervous system activity by capturing the electrical impulses traveling along the postganglionic fibers of the sympathetic nerves to the skin during sympathetic activation. Since myopotential signals, SNA, and noise are all stochastic in nature, separating SNA from ξ(*t*) presents a significant challenge without additional information. To simplify the discussion, we assume that noise is negligible and do not differentiate between SNA and myopotential, particularly for components below 400 Hz. Consequently, we treat ξ(*t*) as SNA. Furthermore, we assume that ξ(*t*) is locally stationary [29], which allows us to account for its nonstationary nature under a “slowly varying” assumption.

## 3. Proposed S3 Algorithm

The proposed SNA recovery algorithm, S3, is designed to recover SNA and can be viewed as a TS-type algorithm incorporating recently developed OS algorithm, eOptShrink [27], followed by nonlocal median filtering. The overall flow of the algorithm is illustrated in Figure 3. Essentially, S3 first recovers the ECG signal and then subtracts the recovered ECG signal to obtain the desired SNA. The algorithm is outlined in detail step by step below.

**Figure 3.**
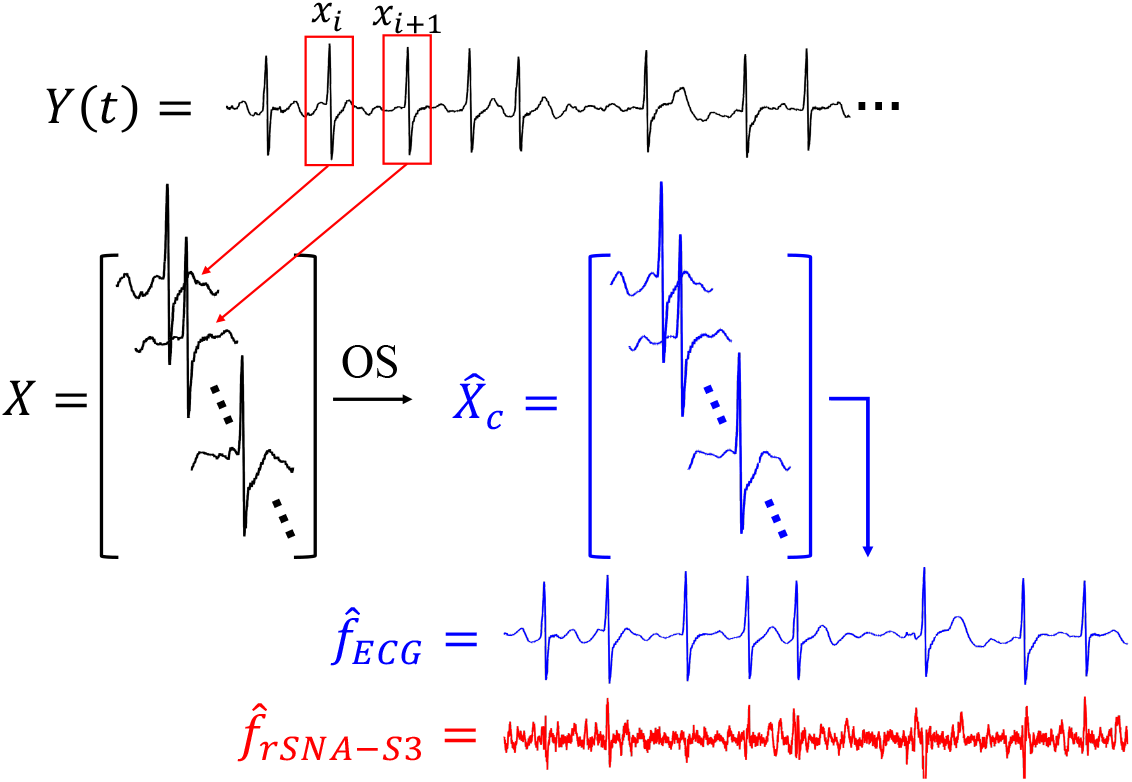
A summary flowchart of the proposed S3 algorithm.

Step 1: Detect cardiac cycle

Denote the input SKNA-I signal as *f*_0_ ∈ ℝ^*N*^, which is sampled at *F*_*s*_ Hz; that is, the sampling period is 1*AF*_*s*_ second. Remove the baseline wandering by applying a high pass filter at the cut 0*V*05Hz, and denote the result *f* ∈ ℝ^*N*^. Apply the standard R peak detection algorithm and let 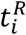 represent the R peak timing of the *i*th cardiac cycle. Suppose *m* intact cardiac cycles are obtained. If the subject is undergoing pacemaker treatment, detect the peaks associated with the atrial and ventricular pacing in DDD mode, denoted as 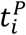 and 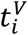 respectively, by identifying peaks in the finite difference of *f*. For VVI mode, following the same procedure to detect the peaks associated with ventricular pacing, denoted as 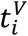.

Step 2: recover the pacing artifacts

This step applies exclusively to subjects undergoing pacemaker treatment. For subjects without a pacemaker, proceed directly to Step 3 by setting *f*_1_ = *f*. If there is atrial pacing, segment the SKNA-I signal into pieces of length 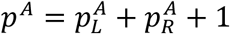, where 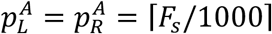 and ⌈*x*⌉ means the smallest integer larger than or equal to *x* > 0, and save the *i*-th cardiac cycle as a *p*^*A*^-dim vector

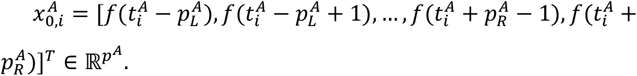

The number *p*^*A*^ depends on the sampling rate *F*_*s*_. Then, remove the linear trend from 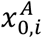 by applying a linear regression, and obtain the atrial pacing artifact, denoted as 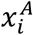. Construct the *atrial pacing matrix* as

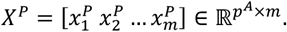

Note that by construction, the 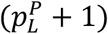 th row of *X*^*P*^ is associated with the peak of the atrial pacing artifact at 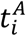.

If there is a ventricular pacing, repeat the same procedure and construct the ventricular pacing matrix 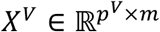 with 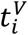, 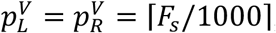, that is, 1*/*1000 second, and 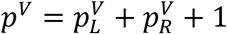. It is important to highlight that *X*^*P*^ represents the sum of the pacing artifact and SNA, as does *X*^*V*^.

Then, apply the matrix denoising step, which is detailed below in the *Key Step*, to reconstruct clean atrial and ventricular pacing artifacts. Denote the resulting clean atrial pacing artifact as 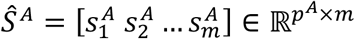 and the clean ventricular pacing artifact as 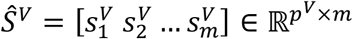. Last, remove the atrial and ventricular pacing artifacts from the SKNA-I signal in the following way. Construct a new signal *f*_1_ ∈ ℝ^*N*^ by setting

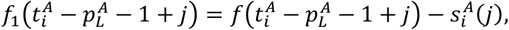

where *i* = 1, …, *m* and *j* = 1, …, *p*^*A*^,

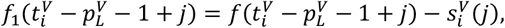

where *i* = 1, …, *m* and *j* = 1, …, *p*^*V*^, and set *f*_1_(*k*) = *f*(*k*) otherwise. In other words, we subtract the clean atrial and ventricular pacing artifacts from *f*.

Step 3: recover ECG

For all cases, construct the ECG data matrix in the following way. Set *p*_1_ and *p*_2_ to be the rounded integer of one half and three quoters of 95% percentile of RR intervals respectively. Segment *f*_1_ (or *f*_1_ = *f* if the subject does not receive a pacemaker treatment) into pieces of length *p* = *p*_1_ *s p*_2_ *s* 1, with the center of R peaks, and save the *i*-th cardiac cycle as a *p*-dim vector

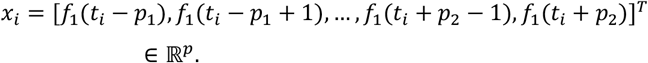

It is possible that *t*_*i*_ − *p*_1_ > *t*_*i*-1_ + *p*_2_ ; that is, two segments of cardiac signal do not overlap, which may happen, for example, when arrhythmia exists. We divide such segment into ⌊(*t*_*i*_ − *t*_*i-*1_ − *p +* 1)*/p*⌋ *+* 1 equally distanced overlapping segments. Suppose in total we obtain *n* segments. Construct an ECG data matrix

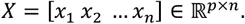

Note that by construction, the (*p*_1_ *s* 1)th row of *X* is associated with the R peaks. Again, *p* depends on the sampling rate.

Then, apply the matrix denoising step detailed below in the *Key Step* to *X* and obtain 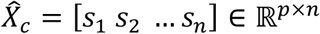 ; that is, *s*_*i*_ is the clean *i* th cardiac cycle after removing SNA. Finally, we recover the ECG signal by stitching all denoised cardiac cycles in 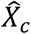 with the overlap-and-add method and denote the resulting denoised ECG signal as 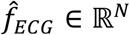.

For subjects undergoing pacemaker treatment, combine the recovered atrial and ventricular pacing artifacts to obtain the pacemaker signal 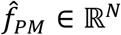. Otherwise, set 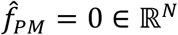.

The remaining part,

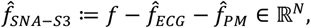

is the desired estimate of SNA, rSNA-S3.

Key Step: Denoise the matrix by optimal shrinkage

This key step is applied to all data matrices referenced above, including *X*^*P*^ and *X*^*V*^ for subjects with pacemakers, as well as *X* for all subjects. To simplify the discussion, consider the ECG data matrix *X* ∈ ℝ^*p×n*^ below and the same procedure applies to other data matrices. Here, *X* represents the summation of the cardiac activity data matrix *X*_*c*_ as a low rank matrix and the SNA matrix *X*_>_. Since the objective is to recover *X*_*c*_, *X*_>_ associated with SNA is treated as “noise”, making *X* a noisy matrix. Recovering the cardiac activity then becomes a denoising problem, for which we apply eOptShrink [27].

Without loss of generality, assume *p* ≥ *n* and set *β*_*n*_ ≔ *pAn*. If *p* < *n*, we simply transpose *X* and apply the same procedure. Also, to simplify the explanation of each step of the algorithm, we assume all entries of *X*_>_ are sub-Gaussian, and refer readers to [27] for the statements under more general setup. Denote the singular value decomposition (SVD) of *X* as 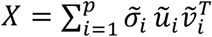, where 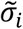 is the singular value and ũ_*i*_ and 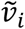 are the associated left and right singular vectors. We assume the singular values are in the decreasing order. Denote 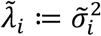, which is the eigenvalues of *XX*^*T*^.

With the above preparation, eOptShrink is carried out by the following three steps. First, evaluate

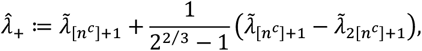

where 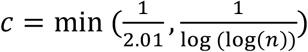 and [*n*^*c*^] is the closest integer to 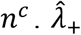 essentially provides an estimate of the noise “strength”. This allows us to determine when the information contained in a singular value can be considered part of the clean data of interest. With 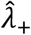, we could estimate the *effective rank* of the noisy dataset, denoted as 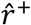, by the number of 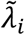 greater than 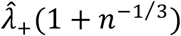 (see Theorems 4.1 and 4.2 in [27]). We shall mention that when a nonzero singular value is not strong enough, with high probability it cannot be detected (see Theorem 3.2 in [27]); in other word, 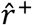 can be strictly less than the rank of *X*_*c*_.

Second, correct small singular values by setting

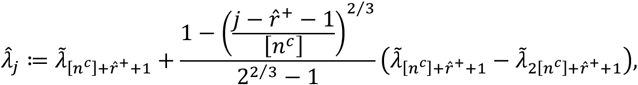

Where 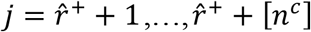*V* Essentially, 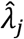is an estimate of the *j* -th eigenvalue of the covariance matrix associated with *X*_>_, which leads to a more accurate noise structure estimate. Second, for 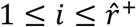, construct 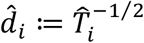, where

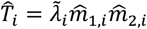

is the numerical implementation of the D transform associated with *X*_>_ (see (15) and (16) in [27]), and

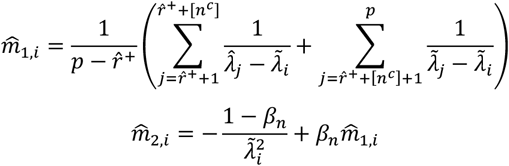

are numerical implementations of Stieltjes transforms associated with *X*_>_.

Next, construct 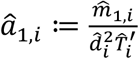 and 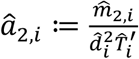, where

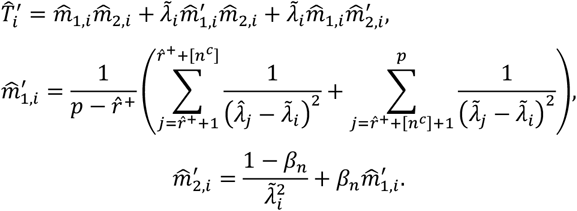

Here, 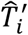, 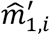 and 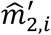are numerical implementations of derivatives of the D transform and Stieltjes transforms associated with *X*_>_. Mathematically, â_1,*i*_ (â_2,*i*_ respectively) approximates the inner products between the true left (right respectively) singular vectors and noisy left (right respectively) singular vectors (see the statement below Proposition 2.6, (28) and Theorem 3.5 in [27]). These seemingly complicated estimates essentially give an estimate of the underlying noise structure via estimating the relationship between the clean singular values and vectors and the noisy singular values and vectors. With these quantities, the last step is denoising the matrix *X* via the formula

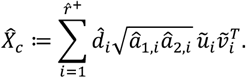

Here, 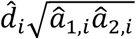 is called the OS of the singular value 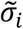, which essentially corrects the noisy singular value 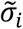 by taking the bias of the singular vectors into account.

To ensure the performance of eOptShrink, we assume that *X*_*c*_ is of low rank, with the rank *r* fixed and much smaller than *p*, and *X*_>_ has a separable covariance structure; that is, *X*_>_ = *A*^1*/*2^Ξ*B*^1*/*2^, where *A* and *B* are positively definite and Ξ contains independent entries with mild moment conditions. The key mathematical fact underlying eOptShrink is that when *β*_*n*_ > 0, the singular values and vectors of *X* are almost surely biased compared to those of *X*_*c*_ as *n* goes to infinite. To our knowledge, there is currently no method available to correct the biased singular vectors. OS accounts for this by optimally correcting the singular values. It has been shown in [27] that 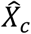 optimally recovers *X*_*c*_ in terms of minimizing the Frobenies norm as the cost function.

We shall comment on these assumptions from practical perspective. Regarding the low rank assumption of *X*_*c*_, recall the geometric meaning of SVD. The left singular vectors determined by SVD can be viewed as an data-driven basis of cardiac cycles and their role is similar to that of Fourier basis commonly used in signal processing. The right singular vectors can be viewed as the “local” coefficients associated with the adaptive basis of cardiac cycles, while the singular values are viewed as the “global” strength of the associated singular vectors. The possibility of imposing this low rank structure comes from two facts. First, the electrophysiological dynamics reflected as P-QRS-T morphology of a cardiac cycle does not change randomly from one to another, and this variation follows some physiological law that we do not specifically quantify. Second, due to the sensitivity of SVD to phase shift, aligning cardiac cycles with R peaks, the most dominant component of the cardiac cycles enhances this similarity and hence enhances the low rank structure. With this geometric meaning in mind, the key step of the proposed S3 algorithm is well aligning R peaks. The separable covariance structure of *X*_>_ captures the short-range and long-range dependence of SNA; that is, *A* captures the short-range dependence of SNA and *B* captures the long-range dependence of SNA. See [27] for more discussion of how this noise matrix impacts the spectrum of *X*.

## 4. Material and statistics

*Material*. We evaluate and demonstrate the performance of the proposed S3 algorithm using one semi-real simulated database, one human database and one mice database.

The semi-real simulated database is constructed as follows. First, for an SKNA-I recording recorded from a healthy subject using ME6000 Biomonitor System (Mega Electronics Ltd, Kuopio, Finland) at a sampling rate of 10,000 Hz, and apply a BPF with spectral cuts at 0.5 to 250Hz to obtain a noisy ECG signal, denoted as *f*_*ECG*,0_. Perform signal quality evaluation to reject low-quality segments, retaining *K* high-quality segments. When *K* >1, for the *k*th high-quality segment, let *t*_*k*_ denote the first R peak and *s*_*k*_ the last R peak. Concatenate the high-quality segments by removing the interval *s*_*k*-1_ and *t*_*k*_ from *f*_*ECG*,)_, and denote the resulting signal as *f*_*ECG*,1_. Next, detect R peaks, and apply the nonlocal median filter with 60 nearest neighbors to obtain a clean ECG, denoted as *f*_*ECG*_. Repeat this procedure to generate 5 clean semi-real simulated ECG signals from 5 SKNA-I recordings. To simulate SNA, we take SKNA-R recorded from a healthy subject using ME6000 Biomonitor System (Mega Electronics Ltd, Kuopio, Finland) at a sampling rate of 10,000 Hz, with bipolar electrodes both placed on the right arm. Sections corresponding to QRS-T segments are discarded, and the remaining portions are concatenated to produce a semi-real SNA signal. Repeat this procedure to generate 5 semi-real simulated SNA signals from 5 SKNA-R recordings. The semi-real SKNA-I recordings are generated by summing the simulated ECG and SNA signals, resulting in a database of 25 recording, each approximately 15 minutes long.

The human database consists of 3 cases, 1 of them are normal subjects, one with Af attack, and one with pacemaker installed and turned on with different pacing modes. We utilized a ME6000 Biomonitor System (Mega Electronics Ltd, Kuopio, Finland) for data acquisition with the sampling rate 10,000 Hz from the first pair of bipolar electrodes placed in the right and left subclavian areas, which is ECG Lead I and denoted as SKNA-I. The data was recorded in the clinical setting and the length is 15 minutes.

The mice database consists of 5 normal mices. We used Tungsten electrode in a lead I configuration and the BIOPAC MP36, a full-featured 4-channel physiological recording system (Biopac Student Lab (BSL) version 4.1), with the sampling rate 10,000 Hz to record SCNA-I. The mices were put in the Faraday cage during the recording to block external electrical fields. The SCNA-I recording was made during baseline, cold pressor test (CPT) and recovery phases (3 minutes for each phase).

### Statistics

For semi-real simulated database, the normalized root mean square error (NRMSE) of the recovered SNA (rSNA) is evaluated, where NRMSE is defined as 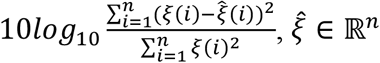 is the recovered SNA and ξ ∈ ℝ^*n*^ is the true SNA. In addition, we evaluate NRMSE_*I*_, where *I* is the spectral band of interest, which is defined as 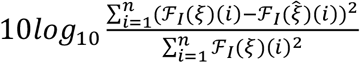, where *ℱ*_1_ means the BPF with the spectral band *I*. We have interest in *I* = [50, 500], [500, 1000], [1000, *T*000] and [*T*000, 5000].

To further assess the performance of S3, we evaluate how well the SNA strength, or sympathetic tone, can be recovered. First, calculate the band-pass filtered (BPF) true SNA, the BPF simulated SKNA-I, and the BPF rSNA-S3. Three BPF ranges are considered: the low-frequency range of 50-300 Hz, which has not been previously addressed in the literature; the spectral range of 300-500 Hz, which was recommended for SNA evaluation using Holter ECG systems; and the widely used spectral range of 500-1000 Hz. The SNA strength is measured by computing the L1 norm of the rSNA over 1-minute intervals, which involves rectifying the rSNA and then calculating the Riemann sum. Then, consider the ratio of strengths between the BPF simulated SKNA-I and the BPF true SNA, called R-BPF, and the ratio of strengths between the BPF rSNA-S3 and BPF true SNA, called R-S3. A ratio closer to 1 indicates better algorithm performance over the given spectral range.

For the real data, since we do not have the true SNA signal, we consider the following *artifact residue (AR)* index. For the *i*-th cardiac cycle, define the *AR*_*i*_ as

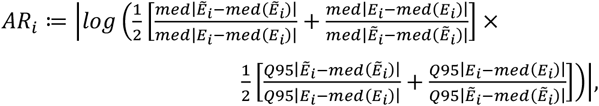

where 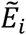 is the rSNA recovered over the ith QRST cycle, *E*_*i*_ is the concatenation of true SNA over the isoelectric period from *i* − 30, …, *i s* 30 cardiac cycles, med means median, Q95 means the 95% quartile, and max means the maximal value. We peak the 10-millisecond interval preceding the onset of the P-wave as the isoelectric point for human database, with the onset of the P-wave identified using a wavelet-based algorithm described in [36]. The algorithm can be downloaded at https://github.com/marianux/ecg-kit. For the mice database, due to the faster heart rate, the isoelectric is selected as the 5-millisecond interval preceding the onset of the P-wave.

The AR index consists of two terms, each capturing distinct statistical properties of the algorithm. The first term assesses how effectively SNA is recovered overall, preventing over-smoothing by comparing the median absolute deviations of the rSNA and the true SNA during the isoelectric period. The second term emphasizes the proper handling of cardiac activity. If the cardiac activity is either inadequately removed or excessively suppressed, the 95th percentile of the rSNA will deviate from that of the SNA during the isoelectric period, causing the second term to diverge from 1. Together, these terms evaluate both the accuracy of cardiac activity removal and the quality of SNA recovery. For an effective reconstruction algorithm, the AR index should be close to 0.

### Comparison

To the best of our knowledge, no algorithm other than BPF specifically focuses on recovering SNA from SKNA-I or SCNA-I. To demonstrate the advantages of the proposed S3 algorithm, we also consider several algorithms originally designed for other types of ECG-related signals but potentially applicable to SNA recovery. These include various implementations of TS, including the single adaptive gain, denoted as TScerutti [30], adaptive gains for P-QRS-T, denoted as TSsuzanna [31] and linear prediction, denoted as TSlp [32], following the nomination in the implementation of these methods [24], the least mean square (LMS) [33], the recursive least square (RLS) [34], the echo state neural network (ESN) [34], and extended Kalman filter (EKF) [35]. The implementation of these methods are available online via [24]. Henceforth, we will systematically use the notation rSNA-*algorithm* to refer to the SNA recovered by the specified algorithm.

To evaluate the performance of various algorithms, the Wilcoxon signed-rank test is employed, with *p* < 0.05 indicating statistical significance. The Bonferroni correction is applied to address the issue of multiple comparisons.

## 5. Result

We quantify and compare the performance of different algorithms on three databases. In the subsequent experiments, all computations and statistical analyses were performed on Windows 11 Pro 23H2, utilizing a 2.3 GHz 8 Core Intel Core i9 processor with 32 GB of 2666 GHz DDR4 memory, and Matlab© version 9.14.0.2337262 (R2023a). The Matlab code and semi-real dataset are available upon request.

### Semi-real simulation

For the semi-real simulated database, the signal lengths are 15.04±4.00 minutes. Table 1 presents the results across various NRMSEs and AR indices. The proposed S3 outperforms other algorithms with statistical significance in all indices.

**Table 1.**
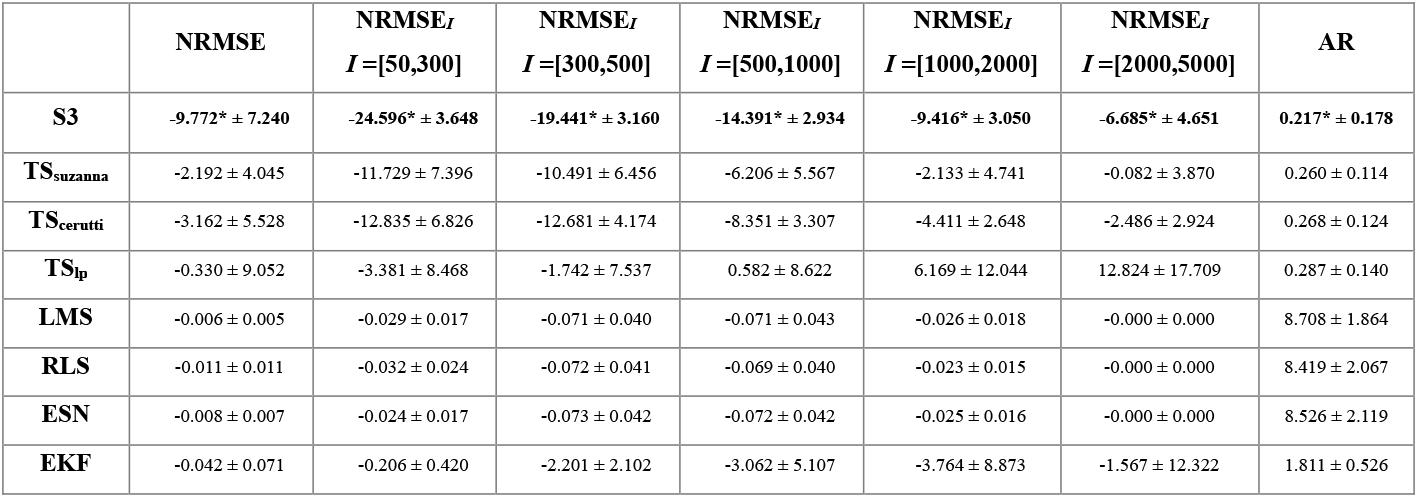
The comparison of the NRMSE and NRMSE_*I*_ of various algorithms applied to the semi-real simulated database, where *I* ⊂ ℝ represents the spectral range of interest. The algorithms being compared are listed in the first column. All indices are reported as mean ± standard deviation. An asterisk next to the mean indicates statistical significance in comparisons between eOptShrink and other algorithms.

Next, consider two groups of signals to quantify how heart rate impacts R-BPF and R-S3. The low heart rate (HR) group consists of the semi-real simulated SKNA-I database, which has an average HR of 1.31 Hz. The high HR group comes from resampling ECG signal in the semi-real simulated database so that the average HR is 1.99 Hz, and summed with the simulated SNA signal. When the spectral range is 50-300Hz (300-500Hz and 500-1000Hz respectively), the mean±standard deviation of R-S3 over the high HR group, R-BPF over the high HR group, R-S3 over the low HR group, and R-BPF over the low HR group are 1.02±0.05, 1.74±0.62, 1.02±0.05 and 1.30±0.24 (1.06±0.06, 1.10±0.07, 1.04±0.05 and 1.06±0.05, and 1.10±0.07, 1.12±0.07, 1.05±0.05 and 1.06±0.04 respectively). Figure 4 presents summary violin plots comparing R-BPF and R-S3 across different spectral ranges and HR groups. Blue asterisks mark cases where R-S3 shows statistically significant differences compared to R-BPF, where all p-values are less than 10^−10^.

**Figure 4.**
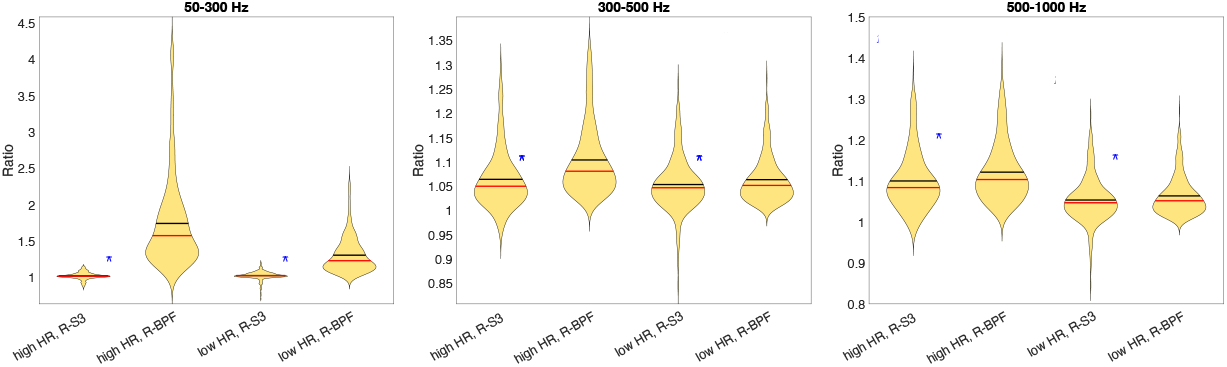
Comparison of SNA recovery over different spectral ranges using the semi-real simulated dataset, where R-BPF and R-S3 are evaluated every 1-minute. From left to right, we show results with the BPF spectral ranges of 50-300 Hz, 300-500 Hz and 500-1000 Hz respectively. The red horizontal line indicates the median and the black horizontal line indicates the mean. Blue asterisks mark cases where R-S3 shows statistically significant differences compared to R-BPF, with all p-values less than 10^−10^.

### Human database

We start with a visual illustration of some results. The first example is from the SKNA-I signal recorded from a subject with an active pacemaker, following a protocol of switching pacing modes: DDD for 6 minutes, VVI for 6 minutes, and DDD for another 6 minutes, with an AV delay of 160 ms. The decomposition results for the VVI mode are shown in Figure 5, while those for the DDD mode are shown in Figure 6. In both figures, the decomposed pacing artifacts, denoted as PM, and ECG are shown in (b) and (c) respectively. The rSNA-S3 is shown in (e), while the rSNA-BPF within the 500-1000 Hz range are shown in (f). (g)–(j) depict the results from other decomposition algorithms. Among the three TS-based algorithms, we include only rSNA-TSlp for visual comparison, as it delivers the most visually satisfactory outcome. It is evident that limited cardiac activity, particularly the QRST complexes, remains in the traditional BPF method within the 500-1000 Hz range, while the traditional BPF method and the EKF approach are unable to eliminate pacing artifacts. ESN and LMS tend to over-filter the signal, while TSlp performs moderately. By contrast, the proposed S3 algorithm effectively decomposes pacing artifacts and ECG signals with high precision, resulting in accurate SNA recovery, although some residual artifacts remain in the DDD mode.

**Figure 5.**
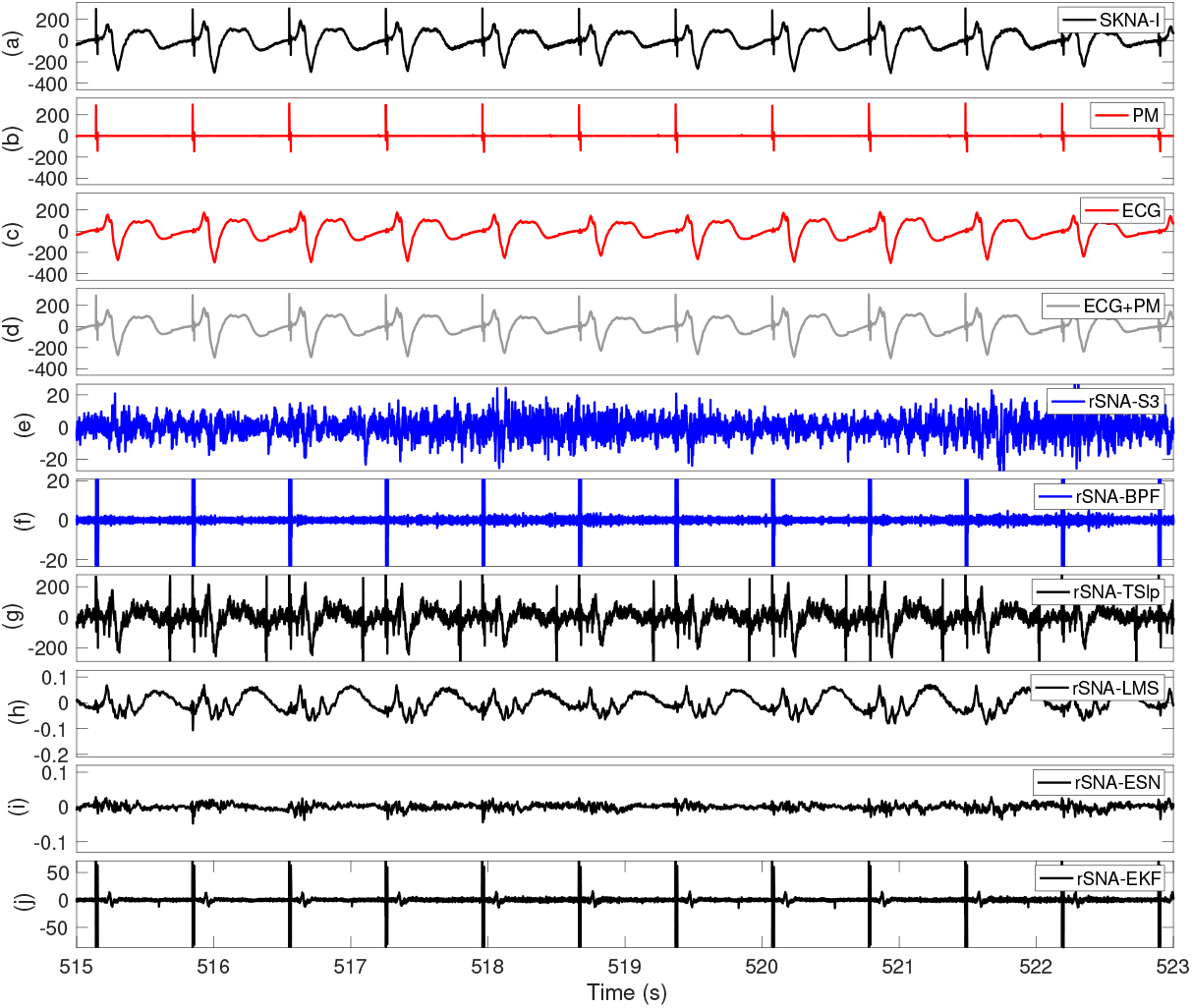
An illustration of the results produced by the proposed S3 algorithm compared to other methods, including the BPF within the spectral range of 500–1000 Hz, where the input data is recorded from a subject with an active pacemaker in VVI mode. From top to bottom, the figure shows: (a) the recorded SKNA-I signal, (b) the pacemaker artifact recovered by the S3 algorithm, (c) the ECG signal recovered by the S3 algorithm, (d) the combination of the pacemaker artifact and the recovered ECG from the S3 algorithm, (e) rSNA-S3, (f) rSNA-BPF, (g) rSNA-TSlp, (h) rSNA-LMS, (i) rSNA-ESN, and (j) rSNA-EKF.

**Figure 6.**
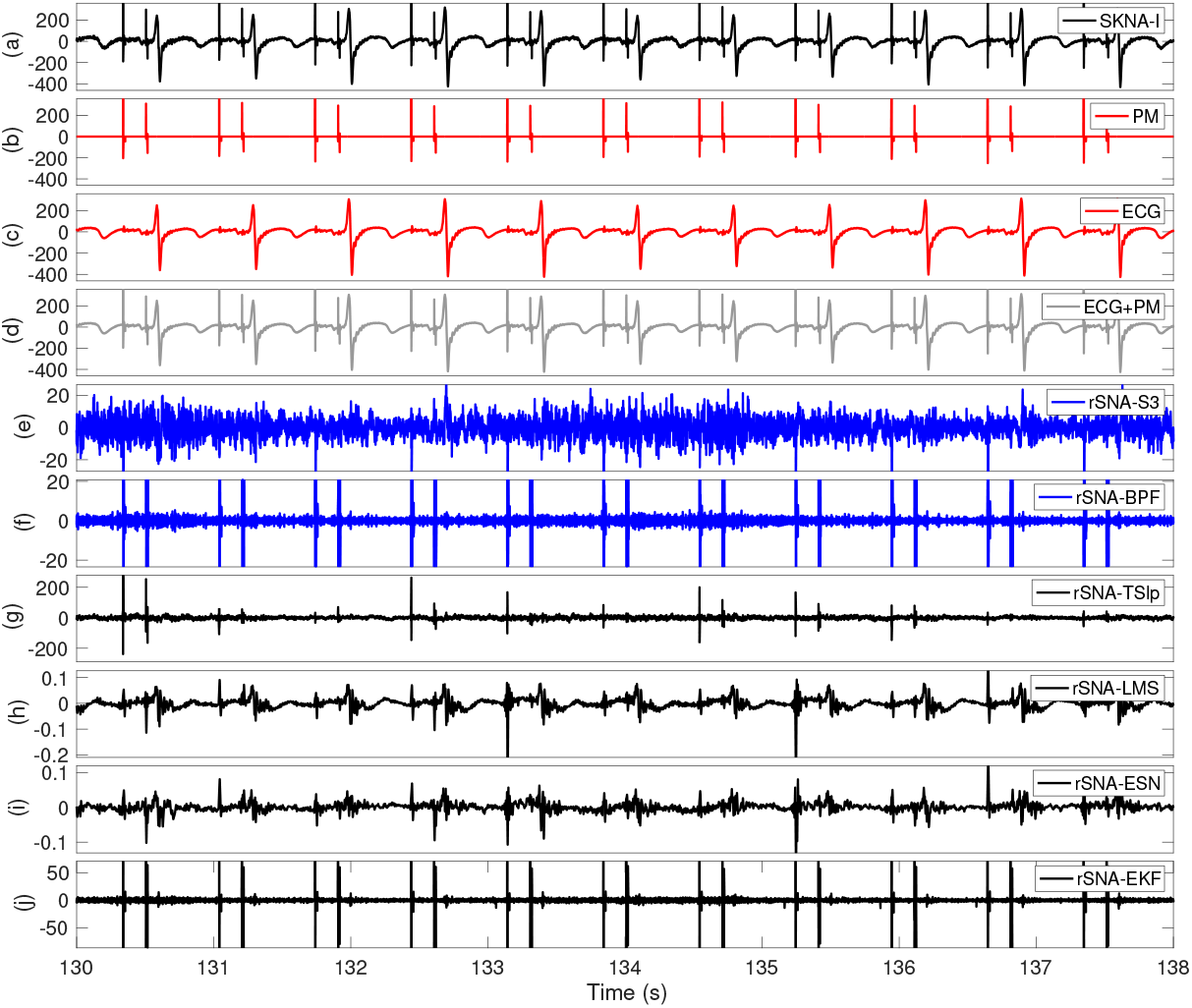
An illustration of the results produced by the proposed S3 algorithm compared to other methods, including the BPF within the spectral range of 500–1000 Hz, where the input data is recorded from a subject with an active pacemaker in DDD mode. From top to bottom, the figure shows: (a) the recorded SKNA-I signal, (b) the pacemaker artifact recovered by the S3 algorithm, (c) the ECG signal recovered by the S3 algorithm, (d) the combination of the pacemaker artifact and the recovered ECG from the S3 algorithm, (e) rSNA-S3, (f) rSNA-BPF, (g) rSNA-TSlp, (h) rSNA-LMS, (i) rSNA-ESN, and (j) rSNA-EKF.

The next example is from the SKNA-I signal recorded from a subject with atrial fibrillation (Af) attack. The decomposition results of different algorithms are shown in Figure 7. The recovered ECG and rSNA-S3 are shown in (b) and (c), while the results of the BPF within the 500-1000 Hz range and other algorithms are shown in (d)-(h). The cardiac activity, particularly the QRST complexes, has been well removed by S3 and the traditional BPF method within the 500-1000 Hz range, while there are still reminding parts in the other methods. Interestingly, EKF can well remove the cardiac activity after 1020^th^ s, but there is still strong artifact remaining during the tachycardia segment before 1020^th^ s. We shall mention that there are some perturbations over the cardiac cycles in the rSNA-S3, which might be artifacts or might have its own clinical significance. See Discussion for more discussion.

**Figure 7.**
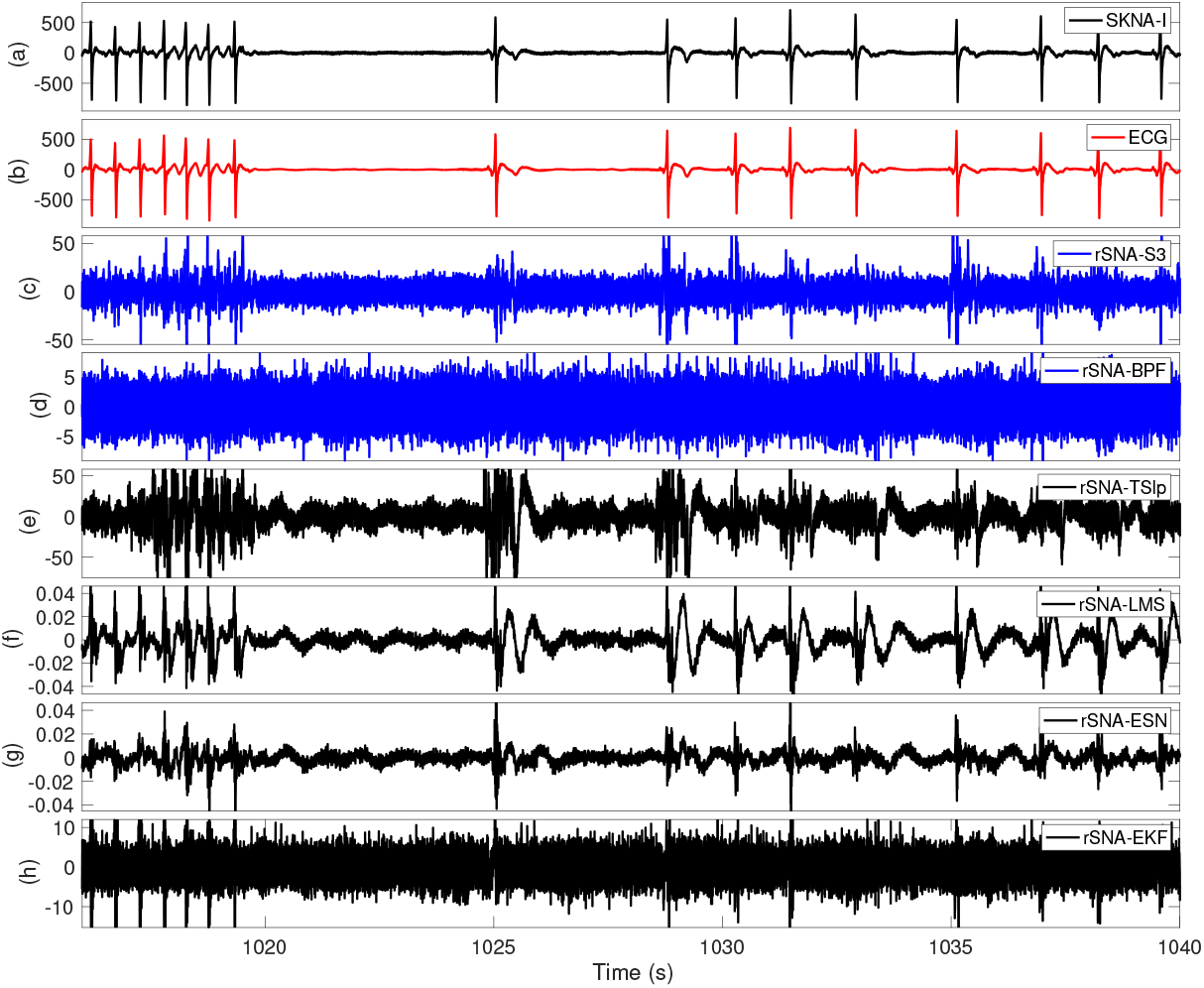
An illustration of the results produced by the proposed S3 algorithm compared to other methods, including the BPF within the spectral range of 500–1000 Hz, where the input data is recorded from a subject with an atrial fibrillation attack. From top to bottom, the figure shows: (a) the recorded SKNA-I signal, (b) the ECG signal recovered by the S3 algorithm, (c) rSNA-S3, (d) rSNA-BPF, (e) rSNA-TSlp, (f) rSNA-LMS, (g) rSNA-ESN, and (h) rSNA-EKF.

Since we do not have the ground truth in the human databases, we utilize the AR index to compare different algorithms. The lengths of signals in the human database are 20.92±1.38 minutes. See Table 2 for a performance comparison of different algorithms, where PM indicates the subject with pacemaker and AF indicates the subject with persistent atrial fibrillation. Overall, the proposed S3 outperforms other algorithms with statistical significance.

**Table 2.**
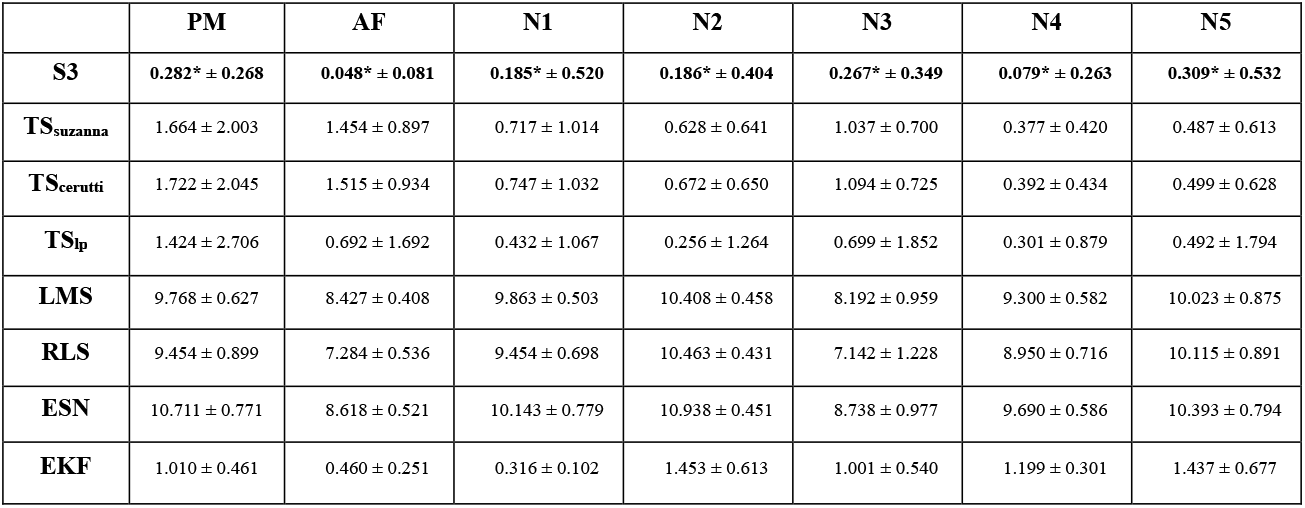
The comparison of Artifact Residue (AR) index of various algorithms applied to the human database. The algorithms being compared are listed in the first column. The AR index is reported as mean ± standard deviation. An asterisk next to the mean indicates statistical significance in comparisons between eOptShrink and other algorithms.

### Mice database

The final analysis features the SCNA-I signal recorded from the mice database, with a result displayed in Figure 8 for visualization. A mouse’s heart rate is typically at least four times faster than that of a human. In this instance, the mouse’s average heart rate is 7.85 Hz. As a result, spectral leakage of cardiac activity into the traditional 500-1000 Hz range is more significant than in the human dataset, making residual cardiac activity visibly evident in the BPF results. In comparison to the S3 algorithm, other methods exhibit clearly noticeable artifacts.

**Figure 8.**
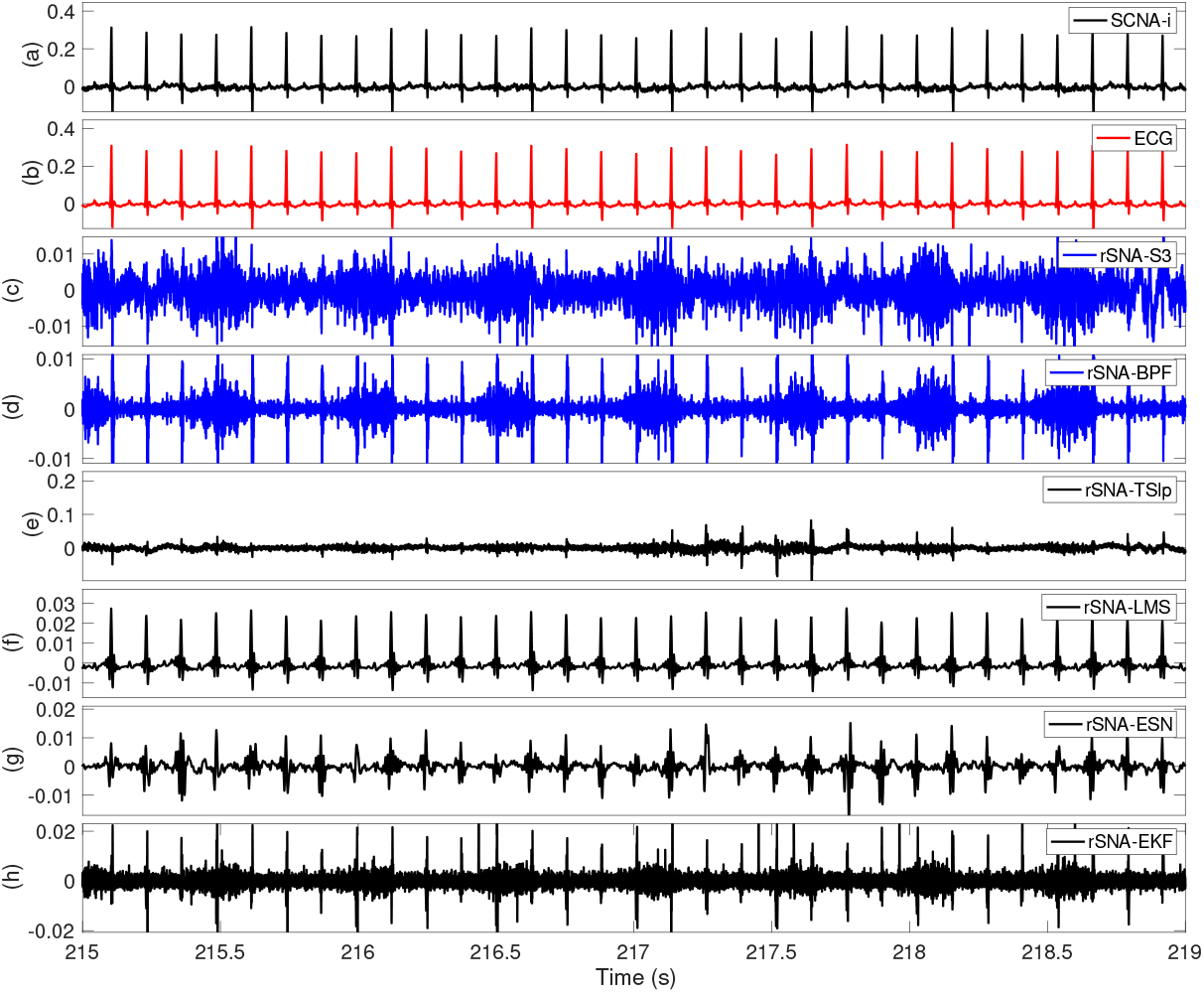
An illustration of the results produced by the proposed S3 algorithm compared to other methods, including the BPF within the spectral range of 500–1000 Hz, where the input data is the SCNA-I recorded from a mice. From top to bottom, the figure shows: (a) the recorded SCNA-I signal, (b) the ECG signal recovered by the S3 algorithm, (c) rSNA-S3, (d) rSNA-BPF, (e) rSNA-TSlp, (f) rSNA-LMS, (g) rSNA-ESN, and (h) rSNA-EKF.

Again, since we do not have the ground truth in the mice database, we utilize the AR index to compare different algorithms. The lengths of signals in the mice databases are 11.88±1.72 minutes. See Table 3 for a performance comparison of different algorithms. Again, overall the proposed S3 outperforms other algorithms with statistical significance.

**Table 3.** The comparison of Artifact Residue (AR) index of various algorithms applied to the mice database. The algorithms being compared are listed in the first column. The AR index is reported as mean ± standard deviation. An asterisk next to the mean indicates statistical significance in comparisons between eOptShrink and other algorithms.

|  | **M1** | **M2** | **M3** | **M4** | **M5** |
| --- | --- | --- | --- | --- | --- |
| **S3** | **0.096\* ± 0.119** | **0.087\* ± 0.183** | **0.208\* ± 0.198** | **0.125\* ± 0.144** | **0.225\* ± 0.293** |
| **TS<sub>suzanna</sub>** | 1.215 ± 0.592 | 0.909 ± 0.611 | 1.506 ± 0.629 | 1.050 ± 1.190 | 1.726 ± 1.123 |
| **TS<sub>cerutti</sub>** | 1.256 ± 0.587 | 0.917 ± 0.611 | 1.602 ± 0.616 | 1.078 ± 1.190 | 1.775 ± 1.073 |
| **TS<sub>lp</sub>** | 0.469 ± 0.554 | 0.230 ± 1.307 | 0.539 ± 0.680 | 0.507 ± 1.098 | 1.320 ± 1.212 |
| **LMS** | 0.656 ± 0.204 | 1.956 ± 0.467 | 0.944 ± 0.308 | 0.611 ± 0.313 | 1.427 ± 0.921 |
| **RLS** | 0.319 ± 0.211 | 1.962 ± 0.475 | 0.928 ± 0.341 | 0.309 ± 0.306 | 1.425 ± 0.936 |
| **ESN** | 0.315 ± 0.217 | 0.787 ± 0.360 | 0.384 ± 0.354 | 0.237 ± 0.251 | 1.056 ± 0.734 |
| **EKF** | 0.108 ± 0.099 | 0.263 ± 0.192 | 0.244 ± 0.161 | 0.181 ± 0.187 | 0.250 ± 0.298 |

### Computational time

From the computational time perspective, it takes 9.84±3.77 (2.33±0.84, 1.89±0.76, 3.37±1.35, 1.38±0.39, 12.14±3.34, 241.73±65.14 and 232.59±65.28) seconds to finish the S3 algorithm (TSsuzanna, TScerutti, TSlp, LMS, RLS, ESN and EKF respectively).

## 6. Discussion

This paper focuses on recovering the SNA signal, and the numerical results demonstrate the applicability of the proposed S3 algorithm. The rSNA signal can be further analyzed for various clinical and scientific purposes. In practice, researchers typically rectify and integrate the rSNA every 100 ms using a Riemann sum to quantify sympathetic tone. This process produces a signal resembling microneurography, which can be displayed over time for visual inspection. In the semi-real simulated database, we show that S3 provides better estimation of sympathetic tone across different spectral ranges, highlighting its potential utility. Additionally, feature extraction using methods like time-varying spectral analysis [36] or time-frequency analysis algorithm like ConceFT [37] could be considered for subsequent analyses.

Notably, S3’s successful recovery of spectral content in the ranges of 50-300 Hz and 300-500 Hz within the semi-real simulated database suggests its applicability to Holter ECG setups [23]. This raises the possibility of convenient, home-based ANS evaluations. However, we cannot claim complete recovery of the SNA signal, especially in the 50-300 Hz range, which includes myopotential interference, since decomposing two potentially dependent random processes remains a challenging problem. Recently, a deep convolutional neural network approach to remove muscle noise was proposed in [38], which might be considered for practical purpose. Addressing this challenge without additional information presents an interesting avenue for future research.

Regardless of the method used, researchers should be aware that similar to the mechanism of ECG-derived respiration [39], rSNA amplitude is influenced by thoracic impedance, which encodes respiratory information. Individual physiological factors such as fluid distribution and cardiopulmonary coupling can also affect rSNA dynamics, impacting clinical applications. These factors must be carefully considered during analysis.

Diseases associated with high sympathetic tone, including heart failure, acute coronary syndrome, cerebrovascular disease, chronic kidney disease, and liver cirrhosis, significantly contribute to morbidity and mortality. In managing these conditions, rSNA from SKNA-I, a simple, non-invasive, and real-time measure of sympathetic tone, has potential clinical utility. Studies have shown the usefulness of rSNA-BPF in managing conditions such as acute coronary syndrome [20], ventricular arrhythmia [12], syncope [18] and overactive bladder [40]. SCNA-I, on the other hand, has been widely applied in basic science and animal study. However, obtaining accurate SNA measurements from SKNA-I or SCNA-I is challenging due to arrhythmia, tachycardia, low sampling rate and various artifacts, as illustrated in Fig. 1. The S3 algorithm is designed to address these challenges and has promising applications in clinical settings.

The relationship between SNA and HRV merits some discussion. While HRV is widely used to assess the ANS in clinical and animal studies, it becomes less reliable in cases of arrhythmias. In contrast, properly recovered rSNA directly measures sympathetic nerve activity and may serve as an alternative for assessing sympathetic function. We hypothesize that rSNA-S3 could assist in developing a new scheme to edit R-to-R interval time series in cases of atrial or ventricular premature contractions [41], a persistent challenge in HRV research.

From a technical perspective, all TS-based algorithms, including S3, rely on accurate cardiac cycle detection, which depends on the signal quality. Despite the availability of advanced R-peak detection algorithms and signal quality assessment methods, occasional missing or erroneous beats still require manual correction. A more accurate R-peak detection algorithm that seamlessly integrates signal quality assessment would enhance the reliability of fully automated systems.

We shall mention another technical point that is essential to explain the limitation of BPF. ECG signals are quasi-periodic, with an average heart rate of approximately 1-2 Hz. Due to the non-sinusoidal nature of ECG waveforms, their spectrum is not confined to 1-2 Hz but extends across a broad range (typically 0.5-150 Hz) due to higher-order harmonics encoding non-sinusoidal patterns such as the P-QRST complexes. These high-frequency harmonics are critical for capturing spike-like features, such as R peaks, and form the basis for the effectiveness of traditional BPF approaches in laboratory setups (500-1000 Hz) or Holter setups (300-500 Hz). However, elevated heart rates, as seen in pediatric patients or animals like mice, shift the spectral content higher. For instance, mice with an average heart rate exceeding 8 Hz exhibit ECG spectral ranges scaling to approximately 4-1200 Hz. Traditional BPFs (e.g., 500-1000 Hz) struggle to isolate cardiac spectral content effectively in such cases, as demonstrated in Figure 8. Similarly, in scenarios involving high heart rates or pacemakers, a BPF range of 300-500 Hz may fail to remove cardiac spectral components. The S3 algorithm addresses these challenges, offering robust performance where traditional BPF methods fall short.

From a theoretical standpoint, the S3 algorithm’s core component, eOptShrink, depends heavily on the noise structure, specifically the SKNA dependence structure as a random process [27]. The separable covariance structure captures both long-range and short-range dependencies in SNA. While this assumption is generally valid, external artifacts or disturbances outside the physiological system may violate it, leading to reduced algorithm performance. Automatically detecting such events and developing a more robust algorithm with theoretical guarantees are essential, particularly for home-based applications. These challenges represent our future research directions.

## 7. Conclusion

We propose a novel algorithm, S3, designed to recover SNA from SKNA-I or SCNA-I signals. The algorithm is grounded in a robust theoretical framework based on random matrix theory and is computationally efficient. Its superior performance across diverse datasets, including semi-real, human, and mice data, from different perspective, including time-domain and frequency domain metrics, demonstrates its advantage over alternative algorithms. We anticipate that S3 will be highly valuable for both clinical and scientific applications.

## Data Availability

The semi-real dataset is available upon reasonable request to the authors

## Acknowledgments

The authors thank Jia-Jhen Lin, Chia-Yun Wang and Wun-Jyun Jhuang for their assistances in the experimental preparation. The authors thank the Center for Laboratory Animals in Kaohsiung Medical University for the animal care.

